# Large-scale integration of omics and electronic health records to identify potential risk protein biomarkers and therapeutic drugs for cancer prevention

**DOI:** 10.1101/2024.05.29.24308170

**Authors:** Qing Li, Qingyuan Song, Zhishan Chen, Jungyoon Choi, Victor Moreno, Jie Ping, Wanqing Wen, Chao Li, Xiang Shu, Jun Yan, Xiao-ou Shu, Qiuyin Cai, Jirong Long, Jeroen R Huyghe, Rish Pai, Stephen B Gruber, Yaohua Yang, Graham Casey, Xusheng Wang, Adetunji T. Toriola, Li Li, Bhuminder Singh, Ken S Lau, Li Zhou, Zichen Zhang, Chong Wu, Ulrike Peters, Wei Zheng, Quan Long, Zhijun Yin, Xingyi Guo

## Abstract

Identifying risk protein targets and their therapeutic drugs is crucial for effective cancer prevention. Here, we conduct integrative and fine-mapping analyses of large genome-wide association studies data for breast, colorectal, lung, ovarian, pancreatic, and prostate cancers, and characterize 710 lead variants independently associated with cancer risk. Through mapping protein quantitative trait loci (pQTL) for these variants using plasma proteomics data from over 75,000 participants, we identify 365 proteins associated with cancer risk. Subsequent colocalization analysis identifies 101 proteins, including 74 not reported in previous studies. We further characterize 36 potential druggable proteins for cancers or other disease indications. Analyzing >3.5 million electronic health records, we conducted analyses of emulated trials for 11 drugs across 290 comparisons and identified three drugs significantly associated with reduced colorectal cancer risk: caffeine vs. paroxetine (HR, 0.51; 95% CI, 0.41–0.64), haloperidol vs. prochlorperazine (HR, 0.47; 95% CI, 0.33–0.68), and trazodone hydrochloride vs. paroxetine (HR, 0.49; 95% CI, 0.38–0.63). Conversely, caffeine was associated with increased cancer risk in comparisons with finasteride (colorectal cancer) and fluoxetine (breast cancer). Meta-analysis identified six drugs significantly associated with cancer risk, including acetazolamide, which was associated with reduced colorectal cancer risk (HR, 0.79; 95% CI, 0.72–0.87). This study identifies novel protein biomarkers and candidate drug targets across six major cancer types and highlights several approved drugs with potential chemopreventive effects.

## Introduction

Human genetic research has not only advanced our understanding of disease mechanisms but has also significantly contributed to drug discovery and development. Drugs supported by genetic evidence exhibit enhanced therapeutic validity compared to those lacking such support, highlighting the importance of incorporating genetic evidence in drug development initiatives^1; 2^. Common risk variants implicated in diseases can dysregulate nearby gene or protein expression, which can mimic the effects of therapeutic drugs on the targetable proteins. These proteins could serve as potential targets for therapeutic intervention^3^. Thus, concerted efforts for cancer prevention based on proteins influenced by common polymorphisms that modulate cancer risk, are urgently needed^4^. To date, genome-wide association studies (GWAS) have identified several hundred common genetic risk loci for each of three prevalent cancer types: breast, colorectal, and prostate^5–8^, and several dozen risk loci have been identified for other cancers, such as cancer of lung, pancreas, and ovarian^9–13^. Previous research, including our work, has identified hundreds of putative cancer susceptible genes potentially regulated by these risk variants, using methods such as expression quantitative trait loci (eQTL) analysis^8–12; 14–20^ and transcriptome-wide association studies (TWAS)^7; 19; 21–29^. However, most dysregulated gene expressions have not been thoroughly investigated at the protein level.

To deepen the understanding of causal mechanisms and enhance drug discovery endeavors, it is imperative to explore data from transcriptomic to proteomic studies. Proteins, the ultimate products of mRNA translation, play critical roles in cellular activities and represent promising therapeutic targets, as evidenced by successful drug targeting of enzymes, transporters, ion channels, and receptors^30^. Recent studies include protein quantitative trait loci (pQTL) mapping and Mendelian randomization (MR) analysis by integrating cancer GWAS and blood proteomics data to identify potential risk proteins. However, only a few dozen of cancer risk proteins have been reported, with a false discovery rate < 0.05^31–36^. Most reported proteins have not been directly linked to the GWAS-identified risk variants in common cancer types. Furthermore, research is lacking in integrating multiple population-scale proteomic studies like the recent emerging UK Biobank Pharma Proteomics Project (UKB-PPP)^37^, which offers an unprecedented opportunity to establish extensive pQTL databases, accelerating therapeutic drug discovery for therapeutic prevention and intervention in human cancers.

Traditional drug discovery faces numerous challenges, including escalating costs, lengthy timelines, and high failure rates^38^. Drug repurposing presents a promising strategy by identifying new applications for existing drugs, leveraging their well-documented characteristics^39^. With the widespread adoption of modern electronic health record (EHR) systems, vast amounts of real-world patient data are available to augment pre-clinical outcomes and facilitate drug repurposing screening. Recently, drug repurposing using EHRs has successfully discovered repurposing hypotheses for preventing Alzheimer’s Disease^40^, reducing cancer mortality^41; 42^, treating COVID-19^43; 44^, and coronary artery disease^45^. However, for therapeutic drugs that have been used for a long term to treat disease indications with evidence of affecting the expression of cancer risk proteins, their potential association with the risk of human cancers remains largely unclear. Some of these drugs may be linked to an increased cancer risk due to long-neglected side effects.

In this work, we integrate large GWAS data for breast, colorectal, lung, ovarian, pancreatic, and prostate cancers and population-scale proteomics data from over 75,000 participants combined from Atherosclerosis Risk in Communities study (ARIC)^46^, deCODE genetics^47^, and UKB-PPP to identify risk proteins associated with each cancer. We further characterized therapeutic drugs based on druggable risk proteins targeted by approved drugs or undergoing clinical trials for cancer treatment or other indications. We further evaluate the effect of cancer risk for those drugs approved for the indications, using over 3.5 million EHR database at Vanderbilt University Medical Center (VUMC). Findings from this study offer novel insights into therapeutic drugs targeting risk proteins for cancer prevention and intervention.

## Material and Methods

### Characterization of lead variants in six types of cancer

We performed a comprehensive analysis to characterize lead genetic variants associated with breast, colorectal, lung, ovary, pancreas and prostate (**Figure 1A**). For breast cancer, we included strong independent association signals at *P* < 1 x 10^-6^ from a fine-mapping study^48^ and risk variants from another GWAS^6^. We combined the reported lead variants from these two studies after removing those variants in LD (*r*^2^ < 0.1 in European populations) (**Figure S1**). Next, we further included additional lead variants from SuSiE fine-mapping analysis on GWAS (N = 247,173)^49^, with fine-mapping windows of 500 kilobases (kb) and allowed a maximum of five causal variants. LD reference was based on the British-ancestry UK Biobank samples (N = 337,000)^50^. We identified a credible set of causal variants with a 95% posterior inclusion probability (95% PIP) for each independent risk signal and a lead variant was represented by the variant with the minimum *P*. We included additional lead variants from our SuSiE analysis with LD *r*^2^ < 0.1 in European populations with the above set of lead variants for those with independent risk-associated signals at GWAS *P* < 5 x 10^-8^ and located in GWAS loci with independent risk-associated signals at *P* < 1 x 10^-6^ in European populations (**Figure S1**).

**Figure 1:**
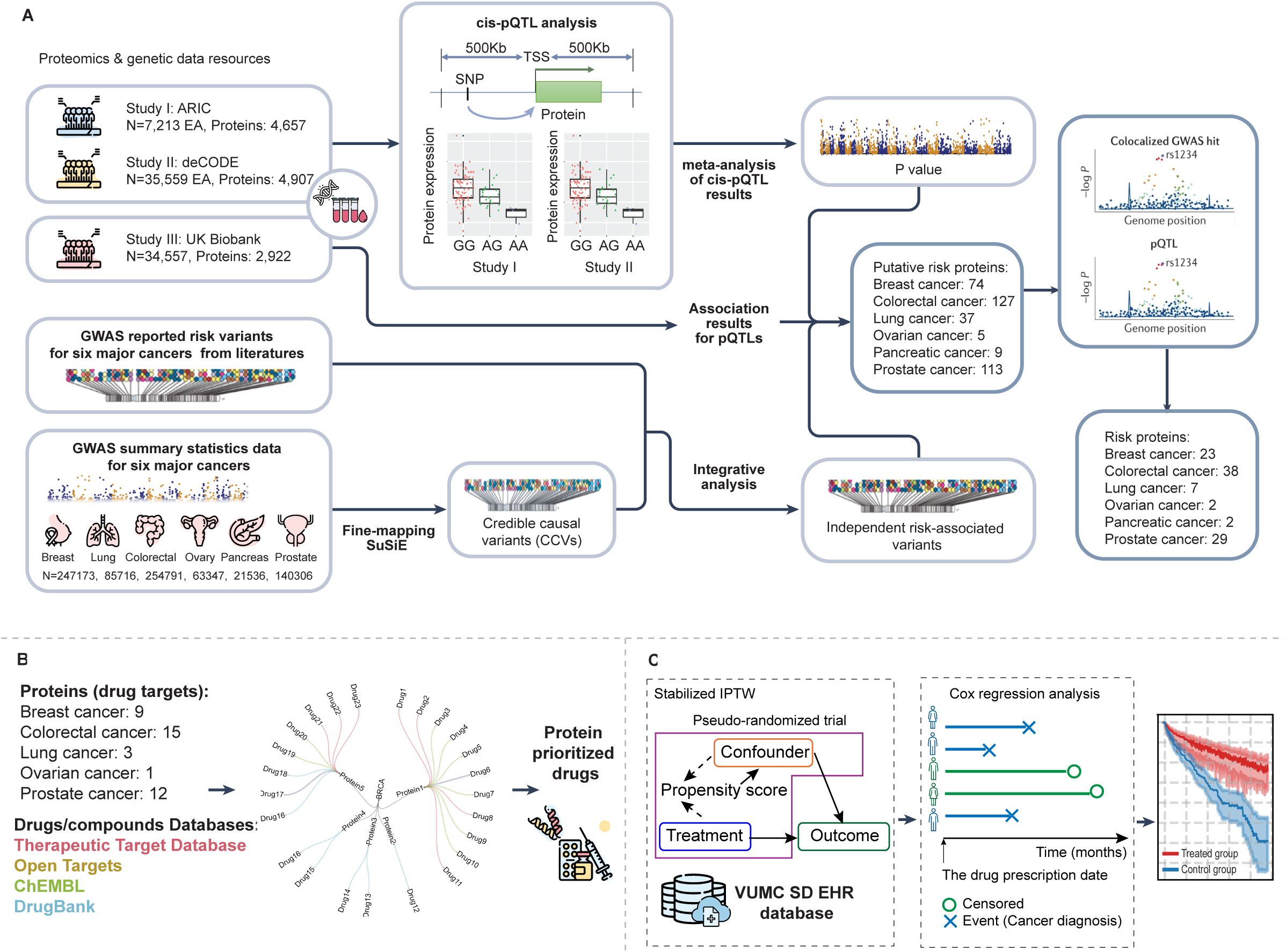
Overview of the Analytical Framework. **A**, An illustration depicting the identification of proteins associated with the risk of the six major cancers: breast, lung, colorectal, ovarian, pancreatic, and prostate. Population-based proteomics data (for pQTLs) and GWAS data resources (for identifying lead variants) utilized in this study are shown in the left panels. Meta-analyses of *cis*-pQTLs from ARIC and deCODE, conducted through the SOMAscan® platform, were combined with pQTL results from the UKB-PPP to identify potential risk proteins, as depicted in the middle panels. Colocalization analyses between GWAS summary statistics and *cis*-pQTLs were performed to identify cancer risk proteins with high confidence, as illustrated in the right panel. **B**, The proteins with evidence of colocalization annotated based on drug-protein information from four databases: DrugBank, ChEMBL, TTD, and OpenTargets. **C**, The framework for evaluating the effects of drugs approved for indications on cancer risk. The Inverse Probability of Treatment weighting (IPTW) framework was utilized to construct emulations of treated-control drug trials based on millions of patients’ Electronic Health Records stored at VUMC SD (left pane). In these emulations, the Cox proportional hazard model was conducted for each trial to assess the hazard ratio (HR) of cancer risk between the treated focal drug and the control drug (right panels).

For colorectal cancer, we analyzed lead variants from our recent fine-mapping study^51^ based on the GWAS data from 254,791 participants in both European and Asian populations. We characterized lead variants with independent risk-associated signals at minimal *P* < 1 x 10^-6^ in European populations, from the analysis based on GWAS from trans-ancestry and European populations, respectively (**Figure S1**). For prostate cancer, we first identified lead variants with independent risk-associated signals at GWAS *P* < 5 x 10^-8^ from our SuSiE fine-mapping analysis on GWAS summary statistics (N = 140,306). We also included additional GWAS-identified risk variants with *P* < 1 x 10^-6^ in European populations from the previous trans-ancestry GWAS^8^ and *r*^2^ < 0.1 with any lead variants from the above set in the fine-mapping analysis (**Figure S1**). Similarly, we used the above strategy to characterize lead variants from fine-mapping analysis for ovarian cancer (N = 63,347) and pancreatic cancer (N = 21,536). We next included additional risk variants that are missed in the above set of lead variants from previous GWAS for ovarian^11^ and pancreatic cancer^10^. For lung cancer, we included risk variants at *P* < 1 x 10^-6^ in European populations from the trans-ancestry GWAS (N=85,716)^9^. Further details on lead variant identification are summarized in **Figure S1** and details on GWAS resources can be found in **Table S1** and **Supplemental Methods I.**

### Identification of putative target proteins for lead variants

To identify potential cancer risk proteins, we mapped GWAS lead variants to *cis*-pQTLs (+/- 500Kb region of a gene) results from three studies among European populations: UK Biobank Pharma Proteomics Project^37^, Atherosclerosis Risk in Communities study^44^ and deCODE genetics^45^ (**Figure 1A**). To increase the power of pQTLs, we combined *cis*-pQTLs from the ARIC and the deCODE (both assayed through SOMAscan^®^ platform) via a a fixed-effects meta-analysis using META^52^. *Cis*-pQTLs from the UKB-PPP (assayed through Olink platform) were independently analyzed. In a few cases where the lead variant did not overlap with any *cis*-pQTLs, we substituted it with the correlated variant exhibiting the strongest association signal. Putative cancer risk protein was defined based on pQTL significance at a Bonferroni threshold of *P* < 0.05 (nominal *P* = 2.3 x 10^-5^, corresponding to 2,164 variant-protein tests for UKB-PPP; nominal *P* = 3.8 x 10^-5^, corresponding to 1,322 variant-protein tests for ARIC+deCODE).

### Colocalization analyses between pQTL and GWAS signals

To identify cancer risk proteins, we conducted colocalization analysis using two approaches: Bayesian method *coloc* ^53^ and summary data-based Mendelian Randomization (SMR)^54^. For the SMR approach, a followed HEIDI test is performed on significant SMR results to determine if the colocalized signals can be explained by one single causal variant or by multiple causal variants in the locus. For each protein, SNPs with *P* < 0.5 from GWAS, MAF > 0.01, and within 50 kb of the lead variant were included. To estimate the posterior probability (PP) of colocalization, we utilized the default priors and coloc.abf function. In our study, we particularly focused on the assumption that one genetic variant is simultaneously associated with both two traits, which was quantified by PP.H4. We considered a protein to host one shared causal variant from GWAS and pQTLs if its *coloc* PP.H4 > 0.5. Additionally, we also performed SMR+HEIDI analysis for significant *cis*-pQTL with default parameter settings. Specifically, significant SMR+HEIDI results were defined as a tested locus with Bonferroni-adjusted SMR *P* < 0.05 and HEIDI *P* ≥ 0.05 (no obvious evidence of heterogeneity of estimated effects or linkage). The above analyses were only conducted in European populations, which were available for both GWAS and proteomics in European populations.

### Functional genomic analyses

For our identified cancer risk proteins, we examined their xQTLs, including eQTLs, sQTLs, and apaQTL using the resource from the GTEx (version 8). We collected eQTLs and sQTLs from six normal tissues and whole blood from GTEx studies, and we collected apaQTLs from Li’s work^55^. A nominal *P* value < 0.05 for at least one xQTL in either tissue or blood samples was considered supportive of the pQTL results.

We identified putative regulatory variants in strong linkage disequilibrium (LD) (*r*^2^ > 0.8 in European population) for lead variants with significant colocalization between GWAS and *cis*-pQTL signals. Using the HaploReg tool^56^, we annotated these variants with a variety of epigenetic annotations, including regulatory chromatin states based on DNAse and histone ChIP-Seq from Roadmap Epigenomics Project, histone marks for promoter and enhancer, binding sites of transcription factors, and gene annotation from the GENCODE and RefSeq. We denoted variants as “Proximal” if they overlapped with these functional annotations near the closest target gene. We analyzed a variety of chromatin-chromatin interaction data, from 4D genome^57^, FANTOM5^58^, EnhancerAtlas^59^, and super-enhancer^60^. We examined the overlap between putative regulatory variants and enhancer elements in corresponding cell lines or tissues of these six cancer types. We further determined enhancer-promoter loops after combining these data with ChIP-seq data of the histone modification H3K27ac (an active enhancer mark). We focused on interacted loops in which a fragment overlapped an H3K27ac peak (enhancer-like elements). In contrast, the other fragment overlapped the promoter of a gene (defined as a region of upstream 2kb and downstream 100bp around transcript start site). We denoted variants as “Distal” if they overlapped with these chromatin-chromatin variants.

For our identified cancer risk proteins, we assessed the statistical significance of their differential protein expression between tumor and normal tissue in breast, colorectal, lung, ovarian, and pancreatic cancer samples using data from CPTAC, accessed through the UALCAN website^61; 62^. Similarly, we analyzed their differential gene expression between tumor and normal tissue using data from TCGA, also through the UALCAN website.

### Identification of focal drugs and formulation of drug-treated patient groups

We assessed the therapeutic relevance of cancer risk proteins by mapping them to known drug–target pairs using four major databases: DrugBank^63^, ChEMBL^64^, the Therapeutic Target Database (TTD)^65^ and Open Targets^66^. This approach identified multiple drugs targeting cancer risk proteins, which we refer to as focal drugs (**Figure 1B**). To evaluate their potential impact on cancer development, we conducted comparative analyses between focal drugs and corresponding control drugs. To minimize potential confounding factors associated with a drug prescription, we selected control drugs that belong to the same second-level Anatomical Therapeutic Chemical classification category (ATC-L2) as the focal drug. For each focal drug, we constructed emulated clinical trials, each comprising a treated group (patients prescribed the focal drug) and a control group (patients prescribed a control drug). One focal drug may have multiple trials, depending on the number of potential control drugs belonging to the same ATC-L2 category. After defining focal-control drugs pairs, we selected patients from the Synthetic Derivative (SD), a de-identified electronic health record database of 3.5 million individuals at Vanderbilt University Medical Center (VUMC)^67^. Using strict eligibility criteria (**Supplementary Methods II**), we ensured each trial included at least 500 patients in both the treated and control groups^40^.

### Calculation of overall hazard ratio for cancer risk drug on cancer development risk

To rigorously evaluate the impact of focal drugs on cancer development, we applied the Inverse Probability of Treatment Weighting (IPTW) framework^68^ to reduce confounding and ensure balanced baseline characteristics between the treated and control groups (**Supplementary Methods III, IV**). Within each eligible trial emulation, we used the weighted Cox proportional hazards model to estimate the hazard ratio (HR) for cancer risk, comparing patients exposed to the focal drug versus those exposed to the control drug over a ten-year follow-up period (**Figure 1C**). For each focal drug, we also performed a random-effects meta-analysis across all balanced trials to estimate its overall association with cancer risk (**Supplementary Methods V**). Finally, to assess the robustness of our findings, we conducted sensitivity analyses using a five-year follow-up window and an alternative control drug selection based on third-level Anatomical Therapeutic Chemical (ATC-L3) classifications, which specifies the pharmacological or chemical properties within the therapeutic subgroup defined at ATC-L2.

## Results

### Characterizing lead variants for breast, ovarian, prostate, colorectal, lung, and pancreas cancers

For breast cancer, we included 196 lead variants with independent association signals at loci from a previous fine-mapping study^48^ and additional 32 genetic variants from another GWAS^6^. Our additional fine-mapping analysis using SuSiE^49^ on Breast Cancer Association Consortium GWAS (N = 247,173) resulted in five novel lead variants. After integrating the previous results with new fine-mapping efforts, we identified 227 lead variants with independently associated with cancer risk at each locus. We applied similar strategies to identify lead variants for colorectal^51^ cancer and other cancer types, leveraging both previously published GWASs and our fine-mapping analyses (**Material and Methods**). In total, we identified 710 lead variants, including 227 for breast cancer, 213 for colorectal cancer, 213 for prostate cancer, 26 for lung cancer, 13 for ovarian cancer and 18 for pancreatic cancer (**Table S2**).

### Identifying cancer risk proteins from pQTLs mapping and colocalization analyses

We mapped the 710 lead variants to *cis*-pQTLs to identify cancer risk proteins. At a Bonferroni-corrected *P* < 0.05, we identified a total of 459 pQTL association signals (corresponding 365 proteins after combined proteins unique for each cancer) for 222 lead variants across six cancer types, including 74 for breast, 127 for colorectal, 37 for lung, 5 for ovarian, 9 for pancreatic, and 113 for prostate cancer (**Figure 2; Table S3**). Notably, 312 of the identified proteins (85.4% of 365) among these cancer types have not been reported in previous proteomics-based MR studies^32–36; 69; 70^ (**Table S4**). Furthermore, through analysis of the identified proteins commonly observed in multiple cancers, we found that 60 proteins were commonly observed in at least two of these six cancers (**Figure S2**). In particular, we observed that several well-known cancer-related proteins, such as HLA-A and HLA-E, were linked to lead variants located in major histocompatibility complex (MHC) in breast, colorectal and lung cancers, highlighting the potential role of these proteins in cancer pleiotropy and shared cancer risk mechanisms (**Figure 2**).

**Figure 2:**
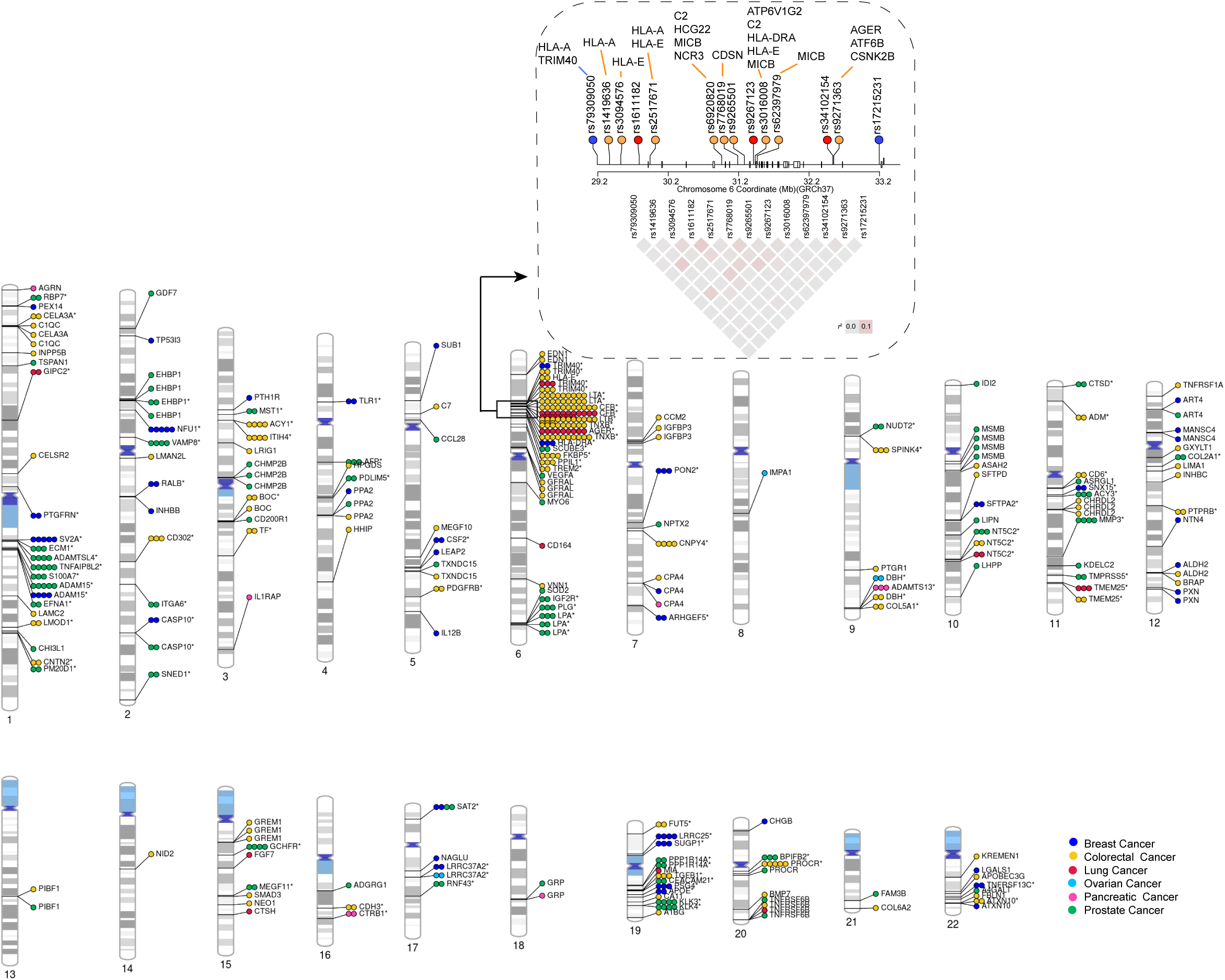
Genome-wide distribution of lead variants and putative risk proteins among six types of cancer. Proteins identified for each cancer are represented by different colors. Each circle represents a single lead variant-protein pair. Proteins marked with an asterisk (*) denote multiple proteins associated with the lead variants. A dashed box highlights several well-known cancer-related proteins, such as HLA-A and HLA-E, which are linked to lead variants located in the major histocompatibility complex (MHC).

A further colocalization analysis identified 101 proteins after combined proteins unique for each cancer that showed strong evidence supported by either colocalization or SMR+HEIDI analysis (**Material and Methods**). Specifically, we identified 23 proteins for breast, 38 proteins for colorectal cancer, 7 proteins for lung, 2 for ovarian, 2 for pancreatic and 29 for prostate cancer, respectively (**Figure 3A, 3B**, **Table S5**). Of these, 74 proteins (73.2% of 101) have not been previously linked to cancer risk (**Table S6**). Of note, 71 proteins were only assayed by either SOMAscan^®^ (n=32) or Olink platform (n=39). For the remaining 22 significant proteins commonly assayed, all showed a pQTL significance signal with a minimal nominal *P* < 1 x 10^-5^ in both ARIC+deCODE and UKB-PPP (*r* = 0.66, *P* = 2×10^-4^; **Figure 3C**). In particular, seven proteins were highlighted as cancer-driver proteins^71;72^ and Cancer Gene Census (CGC)^73^, including ALDH2, HLA-A and SUB1 for breast cancer, ALDH2 and HLA-A for colorectal cancer, NT5C2 for lung cancer, and NT5C2, RNF43, TYRO3 and USP28 for prostate cancer.

**Figure 3:**
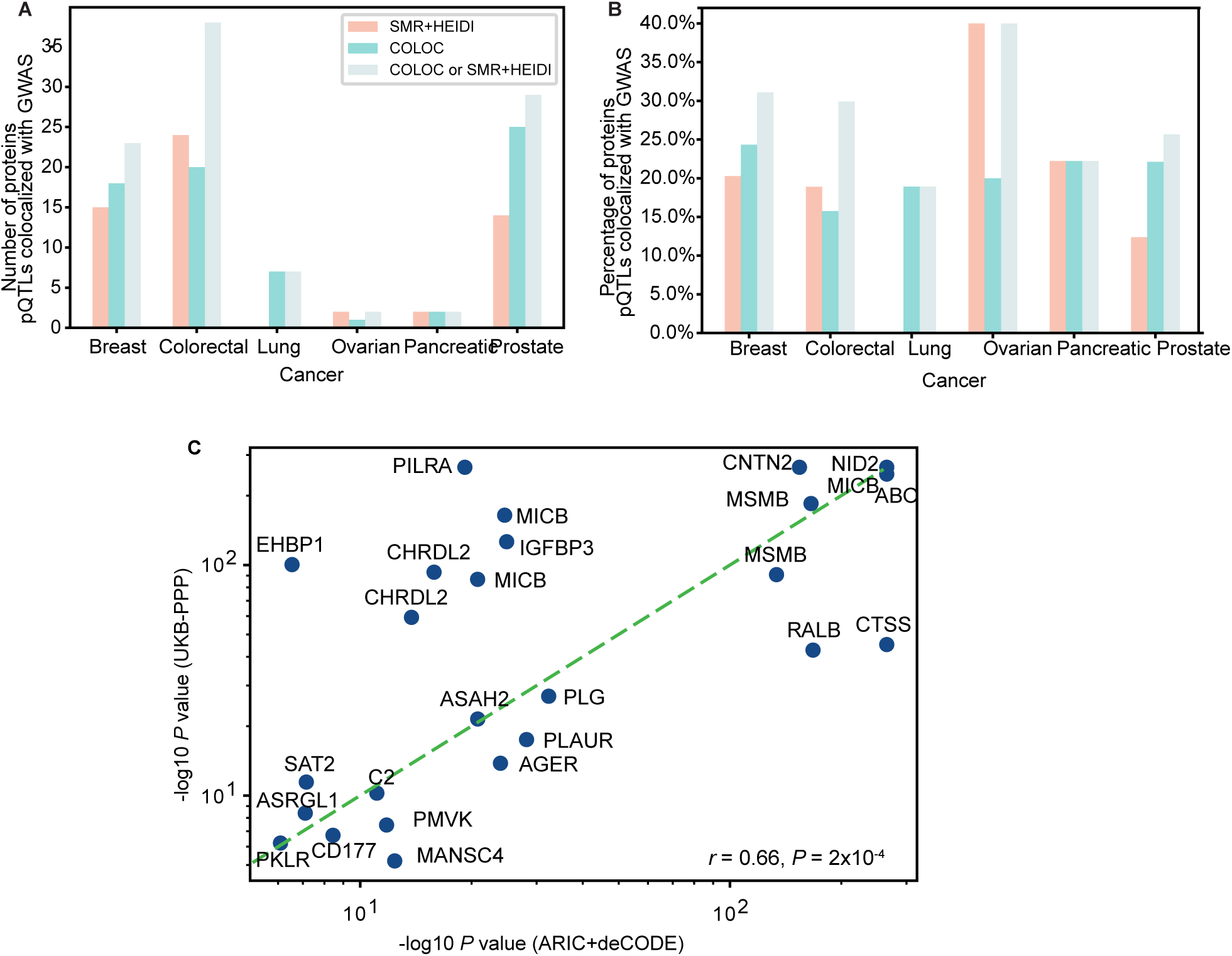
Identification of 101 cancer risk proteins through pQTL and colocalization analysis. **A**, Number of proteins showing evidence of colocalizations between pQTLs and GWAS association signals for six cancer types**. B**, Percentage of proteins showing evidence of colocalizations between pQTLs and GWAS summary statistics for six cancer types. **C,** A plot illustrating the high consistency of pQTL p-values for 22 cancer risk proteins between the ARIC+deCODE and the UKB-PPP (proteins commonly assays from SOMAscan^®^ and Olink platforms).

### Cancer risk proteins supported by functional genomics analyses

Of the identified 101 proteins among the six cancers, we examined whether they are supported by functional genomics analyses. Specifically, we first evaluated xQTL (i.e., eQTLs, alternative splicing - sQTLs, and alternative polyadenylation - apaQTLs) results in their respective target tissues and whole blood samples (**Material and Methods**). We found 63 proteins that were supported by at least one xQTLs at a nominal *P* < 0.05, including12 for breast (52% of 23), 22 for colorectal (57% of 38), 5 for lung (71% of 7), 2 for ovarian, 2 for pancreatic, and 20 (68% of 29) for prostate cancer (**Table S6**). Second, we used functional genomic data generated in their cancer-related tissues/cells (i.e., promoter and enhancer) to characterize putative functional variants that are in strong LD (*r*^2^ > 0.8 in the European population) with the lead variants (**Material and Methods**) Our results showed that 17 genes were likely regulated by the closest putative regulatory variants with either promoter and/or enhancer activities (**Table S7**). We further investigated the potential distal regulatory effects of putative functional variants on these genes by analyzing chromatin interaction data (**Material and Methods**). We found that 39 genes were regulated distally by putative functional variants through long-term promoter-enhancer interactions (**Table S8**). Lastly, we examined differential protein expression between normal and tumor tissues available for breast, colon, lung and pancreatic cancers using data from Clinical Proteomic Tumor Analysis Consortium (CPTAC). We showed evidence of the 18 identified proteins with consistent association directions supported by significantly differential expression at a nominal *P* < 0.05, including 3 for breast cancer, 10 for colorectal cancer, 4 for lung cancer and 1 for pancreatic cancer. Similarly, we showed evidence of the 40 identified proteins supported by significantly differential mRNA expression using data from The Cancer Genome Atlas Program (TCGA), including 6 for breast cancer, 15 for colorectal cancer, 3 for lung cancer, 1 for pancreatic cancer and 15 for prostate cancer. Taken together, our analysis provided additional evidence that most of the identified proteins are partially or wholly supported by functional genomics analyses (**Table S9)**.

### Identifying druggable proteins

Using data from DrugBank^74^, ChEMBL^75^, the Therapeutic Target Database^76^ (TTD) and OpenTargets^77^, we comprehensively annotated our proteins as therapeutic targets of approved or clinical-stage drugs. Of the 101 proteins among the six cancers, we identified 36 druggable proteins potentially targeted by 404 approved drugs or undergoing clinical trials for cancer treatment or other indications (**Figure 4**, **Table S10**). Specifically, we found 19 proteins targeted by 133 drugs either approved or under clinical trials to treat cancers (**Figure 5, Table S11**). Our results also provide evidence that the remaining draggable proteins are targeted by 197 drugs used for treating indications other than cancer (**Figure S3**).

**Figure 4:**
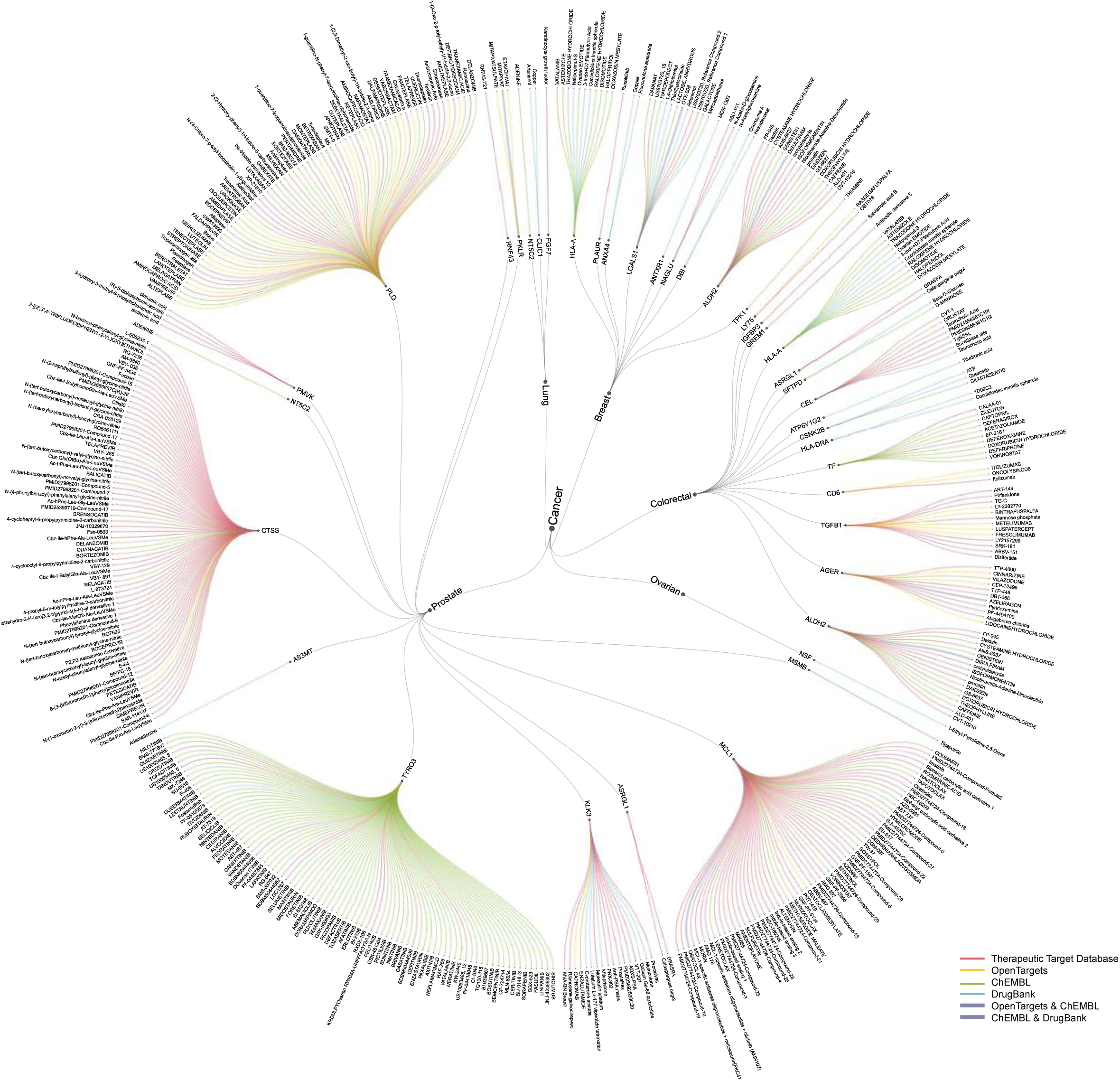
A circular plot showing 36 druggable proteins potentially targeted by 404 approved drugs or undergoing clinical trials for cancer treatment or other indications. Presented from inner to outer layers are cancer types, proteins, and drugs. Each drug-protein interaction is annotated by DrugBank, ChEMBL, TTD, and OpenTargets, with lines in different colors representing each database. Interactions where proteins are annotated by two databases are linked to drugs with thick lines.

**Figure 5:**
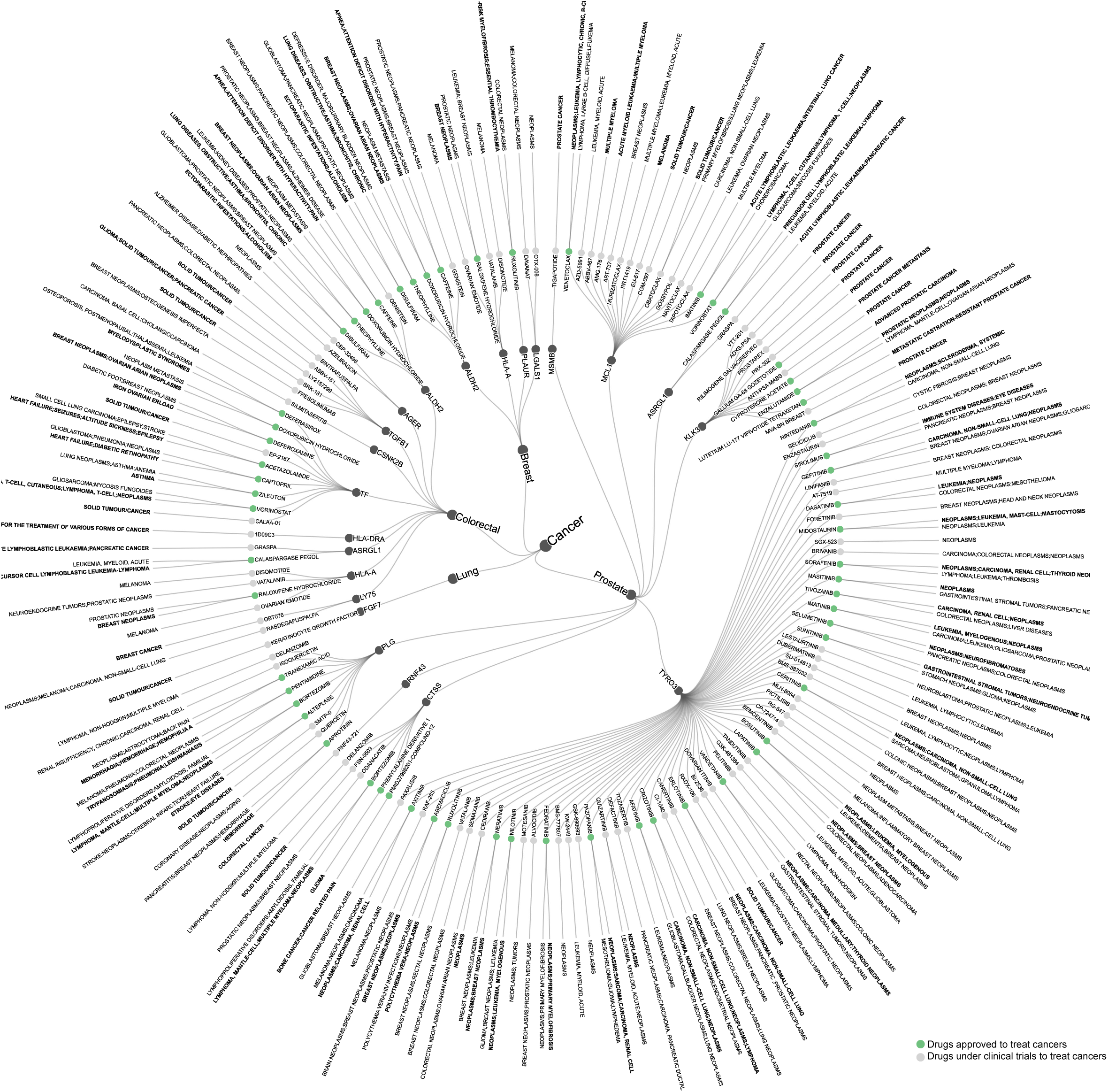
A circular plot showing 19 druggable proteins potentially targeted by 133 approved drugs or undergoing clinical trials for cancer treatment. Presented from inner to outer layers are cancer types, proteins, drugs and cancers. Drugs approved and undergoing clinical trials for cancer treatment are highlighted on green and gray, respectively. Drug approved indications are formatted in bold, while indications under clinical trial are in regular font.

### Evaluating associations of drugs approved for indications with cancer risk

We next evaluated the effect on cancer risk of therapeutic drugs that have been used long-term to treat non-cancer indications based on real-world EHRs from the VUMC Synthetic Derivative (SD) database. Following these stringent criteria descried in **Material and Methods**, we formed 290 balanced trials from 11 treated drugs, associated with seven proteins from three cancers (**Figure 6A**), each with more than 10 eligible control drugs. Specifically, we identified three drugs for breast cancer: Caffeine targeting ALDH2, and Haloperidol and Trazodone Hydrochloride with both targeting HLA-A; several drugs for colorectal cancer: Caffeine and Theophylline with both targeting ALDH2; Acetazolamide and Captopril, with both targeting TF; Haloperidol and Trazodone Hydrochloride, with both targeting HLA-A; and Vilazodone targeting AGER; and one drug for prostate cancer: Sirolimus targeting TYRO3 (**Figure 6B**). All patients analyzed had substantial medical records during the ten-year follow-up period, with a median follow-up of ∼7 years (85 months) and an interquartile range (IQR) of 6-8.5 years (72-102 months). Additionally, across the 290 trials, a median of 99.3% of patients were either cancer-free at the end of the follow-up period (median: 58%) or were lost to follow-up, while the remaining 0.7% were diagnosed with cancer (**Table S13**).

**Figure 6:**
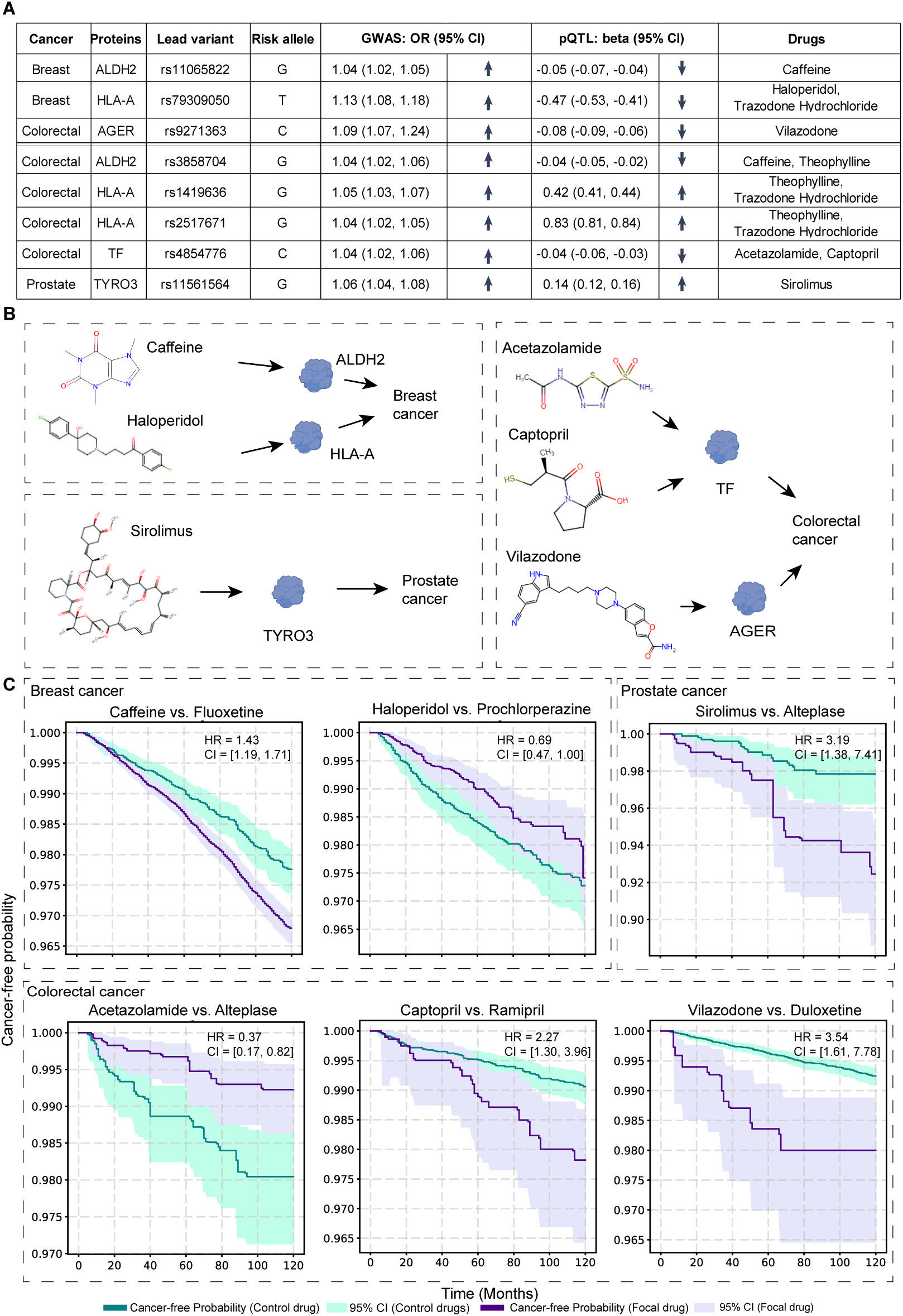
Drugs approved for treated indications showing significant effects on cancer risk. **A**, A table showing cancer risk alleles of lead variants, risk proteins, and drug name approved for indications. Positive associations are indicated by upward arrows, while negative associations are indicated by downward arrows. Odds ratio (OR) refers to the exponential transformation of logistic regression coefficient and beta refers to linear regression coefficient. **B**, An illustration of six drugs, showing significant associations with cancer risk in meta-analysis, and their potential targeted risk proteins. **C**, Survival plots depict the statistically significant difference in the probability of being cancer-free for patients in the treated group (taking a focal drug, shown in green) and those in the control group (primarily selected by the smallest *P* value across eligible treated-control trials for each significant drug in the meta-analyses, shown in purple; all detailed results see Table S13). The shaded area represents the 95% confidence interval. The hazard ratio and *P* for the focal drug compared to the control drug, determined through weighted Cox proportional hazards models, are presented in the top right corner of each panel.

For breast cancer, we applied the weighted Cox proportional hazards model to calculate the hazard ratio (HR) for each trial within the ten-year follow-up period for analyzed three drugs (Caffeine, Haloperidol, and Trazodone Hydrochloride) across 73 eligible treated-control drug trials. Particularly, at a Bonferroni-corrected *P* < 0.05 (*P* < 0.05/290), we found that Caffeine was associated with an increased risk when compared to Fluoxetine (HR = 1.43, 95% CI = [1.19, 1.71]) (**Table S13**). Furthermore, we conducted a random-effects meta-analysis for each of these three drugs. At a Bonferroni-corrected *P* < 0.05 (*P* < 0.05/11), we found that Caffeine was associated with an increased risk (HR = 1.15, 95% CI = [1.09, 1.20]), while Haloperidol was associated with a decreased risk (HR = 0.86, 95% CI = [0.78, 0.95]) (**Table S14, Figure 6C**).

For colorectal cancer, we analyzed seven drugs (Acetazolamide, Caffeine, Captopril, Haloperidol, Theophylline, Trazodone Hydrochloride, and Vilazodone) across 185 eligible treated-control trials. At a Bonferroni-corrected *P* < 0.05, we found that Caffeine was associated with a decreased risk when compared to Paroxetine (HR=0.51, 95% CI = [0.41, 0.64]) and Magnesium Sulfate (HR=0.36, 95% CI = [0.27, 0.48]). Conversely, Caffeine, when compared to the control drug Finasteride (HR=2.32, 95% CI = [1.67, 3.23]), was associated with an increased risk. We also identified that Haloperidol and Trazodone Hydrochloride were associated with an increased risk when compared to the Prochlorperazine (HR = 0.47, 95% CI = [0.33, 0.68]) and Paroxetine (HR = 0.49, 95% CI = [0.38, 0.63]), respectively (**Table S13**). Further meta-analyses showed that Acetazolamide (HR = 0.79, 95% CI = [0.72, 0.87]) was associated with a decreased risk, while Captopril (HR = 2.15, 95% CI = [1.81, 2.57]) and Vilazodone (HR = 1.73, 95% CI = [1.45, 2.05]) were associated with an increased risk (**Table S14, Figure 6C**).

For prostate cancer, we analyzed one drug (Sirolimus) across 32 eligible treated-control trials. At a Bonferroni-corrected *P* < 0.05, none of the trials comparing Sirolimus to its control drugs showed a significant association with cancer risk. However, our meta-analysis revealed that Sirolimus was significantly associated with an increased risk in prostate cancer (HR = 1.37, 95% CI = [1.2, 1.55]) (**Table S14, Figure 6C**).

In addition, we conducted analyses using the weighted Cox proportional hazards model within the five-year follow-up period for the significant treated-control trials identified above, demonstrating consistent associations across all six trials, at a nominal *P* < 0.05 (**Table S15**). Similarly, with meta-analyses based on the results from the five-year follow-up period, we showed five of the identified six drugs (breast cancer: Caffeine; colorectal cancer: Acetazolamide, Captopril and Vilazodone; prostate cancer: Sirolimus) showed consistent associations at a nominal *P* < 0.05 (**Table S14**). Additionally, when meta-analyzing trial emulations at the ATC-L3 level, a narrower pharmacological or therapeutic subgroup, three drugs (colorectal cancer: Captopril and Vilazodone; prostate cancer: Sirolimus) maintained consistent direction of association with cancer risk, as compared to ATC-L2, in both the ten-year and five-year follow-up periods (nominal *P* < 0.05) (**Table S16**).

## Discussion

In this study, we conducted a comprehensive investigation of cancer risk proteins by integrating lead variants and pQTLs for six common cancer types using large-scale GWAS and population-based proteomics data. Through pQTL mapping and subsequent colocalization analysis, we identified 101 risk proteins across the six cancer types, with over three-quarters of them not previously linked to cancer susceptibility. Moreover, most of the proteins we identified are supported by functional genomics analyses. Our findings not only significantly expand the pool of known cancer risk proteins but also offer new insights into the biology and susceptibility of common cancers.

Through analysis of drug-protein interaction databases, we identified 36 druggable proteins potentially targeted by 404 therapeutic drugs. Among these, 30 drugs have already received approval for cancer treatment, while 73 are currently undergoing clinical trials for cancer treatment. These findings offer genetic evidence supporting the effectiveness of certain drugs and suggest potential opportunities for repurposing them to treat additional cancers that share common risk proteins. However, it’s crucial to acknowledge that while the cancer risk proteins identified in our study hold promise as therapeutic targets for cancer treatment, drugs may also have adverse effects, potentially exacerbating cancer development through these targets (i.e., depending on their inhibitory or promotive effects)^78^. Additionally, our analysis characterized 197 drugs used for indications other than cancer, which may influence cancer risk due to their interactions with cancer-risk proteins. Overall, our findings have the potential to accelerate therapeutic drug discovery for the prevention and intervention of human cancers.

Our results showed that Acetazolamide exhibits a notable effect in preventing colorectal cancer development, through targeting the colorectal cancer risk protein, transferrin (TF). Previous literature suggested that TF protein may function similarly to an inhibitor of CA (e.g., ICA protein), as ICA belongs to the same TF protein superfamily^79; 80^. Thus, our genetic discovery provides strong evidence supporting acetazolamide as a promising drug for colorectal cancer prevention, potentially influencing a TF-regulated CA etiological pathway. In line with our findings, prior studies demonstrated Acetazolamide’s role in inhibiting cell viability, migration, and colony formation ability of colorectal cancer cells^81^, as well as its ability to suppress the development of intestinal polyps in Min Mice^82^. Interestingly, we also identified another drug, Captopril, which potentially targets the cancer risk protein TF and is associated with an increased colorectal cancer risk. Captopril may influence cancer risk by inhibiting Angiotensin-converting enzyme (ACE), thereby affecting downstream TF protein levels through the regulation of its receptors^83; 84^. Consistent with our findings, a previous study also highlighted that ACE inhibition may play a risk-promoting role in colorectal cancer^85^.

Our result indicated that Caffeine, a drug prioritized by the risk protein ALDH2, may have a protective role in colorectal cancer when compared to several control drugs. These findings align with previous studies suggesting a potential protective effect of Caffeine in colorectal cancer^86; 87^. Specifically, Caffeine demonstrated a significant protective role in colorectal cancer risk when compared to Paroxetine (*P* = 5.66 x 10^-9^) and Magnesium Sulfate (*P =* 9.10 x 10^-12^). This is supported by previous studies highlighting associations between ALDH2 variants and Caffeine consumption^88; 89^ and another study demonstrating that Caffeine inhibits ALDH2 activity, resulting in reduced nucleophilicity and partial alterations in the enzyme’s secondary structure^90^. Additionally, ALDH2 has been shown to enhance colorectal cancer stemness by activating β-catenin signaling^91; 92^. Conversely, our results also revealed that Caffeine exhibits a risk-promoting role in colorectal cancer when compared to Finasteride (*P =* 4.72 x 10^-7^). This suggested that Finasteride may have a stronger protective effect against colorectal cancer risk than Caffeine, as previous studies have shown that Finasteride may play a role in the causal pathways of colorectal neoplasms^93^ and demonstrated effectiveness in the treatment of prostate neoplasms^94–96^.

Our meta-analysis results showed that Haloperidol, which targets the known cancer risk immune protein, HLA-A, is associated with a reduced breast cancer risk. This contrasts with previous studies suggesting that Haloperidol increases breast cancer risk^78; 97; 98^. The meta-analysis result appears to be primarily driven by the most significant individual trial (Haloperidol vs. Prochlorperazine, HR = 0.69, *P* = 4.72 × 10^−2^), which may overshadow other signals showing the risk-promoting role played by Haloperidol. Interestingly, our meta-analysis results showed that Haloperidol plays a risk-promoting role in colorectal cancer (HR = 1.25, CI = [1.01, 1.54], nominal *P* = 4.0 x 10^−2^). Although these findings are in line with the observation that HLA-A is associated with decreased risk in breast cancer and increased risk in colorectal cancer, replication of these findings in an independent dataset is necessary for future studies.

We also uncovered an additional drug, Vilazodone, which is associated with an increased colorectal cancer risk by targeting AGER, a protein known to promote chronic inflammation by activating the NF-κB pathway^99^. Our study also found Sirolimus, a mechanistic target of rapamycin (mTOR) inhibitor, exhibits a risk-promoting role in prostate cancer, consistent with previous research^100; 101^. While our findings highlight potential etiological pathways involving these drugs, it is essential to evaluate the effects of the reported candidate drugs through both *in vitro* and *in vivo* assays in future research.

## Data Availability

All data produced in the present study are available upon reasonable request to the authors

## Data availability

Table S1 provides the download information for the summary statistics of GWAS data for the six common cancers, including breast, ovary, prostate, colorectum, lung, and pancreas.

Metadata and pQTL summary statistics from UKB-PPP can be downloaded from Synapse: Project SynID: syn51364943; pQTL from ARIC^46^ and deCODE genetics^47^ can be accessed through previous publications (PMID: 34857953 and PMID: 35501419). Functional genomic data includes: TCGA and CPTAC differential expression results accessible through https://ualcan.path.uab.edu/index.html; 4DGenome: https://4dgenome.research.chop.edu/; Depmap : https://depmap.org/portal/; FANTOM5: http://fantom.gsc.riken.jp/5/. HaploReg: https://pubs.broadinstitute.org/mammals/haploreg/.GTEx: https://gtexportal.org/home/. GENCODE (v26.GRCh38) was downloaded from https://www.gencodegenes.org/human/release_26.html. National Cancer Institute can be accessed through https://www.cancer.gov/about-cancer/treatment/drugs; CGC can be accessed accessed via COSMIC website: https://cancer.sanger.ac.uk/census. Drugs and compounds data can be downloaded from the following URLs: ChEMBL: https://www.ebi.ac.uk/chembl/; Therapeutic Target Database: https://db.idrblab.net/ttd/; Open Targets: https://www.opentargets.org/; DrugBank: https://go.drugbank.com/. The EHR data, containing de-identified clinical information, can be accessed through the VUMC SD database. Data is available through restricted access for approved studies and researchers who agree to specific conditions of use.

## Code availability

The developed pipeline and main source R codes that are used in this work are available from the GitHub website of Xingyi Guo’s lab: https://github.com/XingyiGuo/PQTL_EHR/

## Declaration of Interests

The authors declare no competing interests.

## Acknowledgments

This work was supported by the US National Institutes of Health grant 1R37CA227130-01A1 and R01CA269589-01A1 to X.G. The data analyses were conducted using the Advanced Computing Center for Research and Education (ACCRE) at Vanderbilt University. New Frontiers in Research Fund (NFRFE-2018-00748) and NSERC Discovery Grant (RGPIN-2024-04679) to Q.L. The computational infrastructure was partly supported by a Canada Foundation for Innovation JELF grant (36605) to Q.L.

## Supplementary Figures

**Figure S1:**
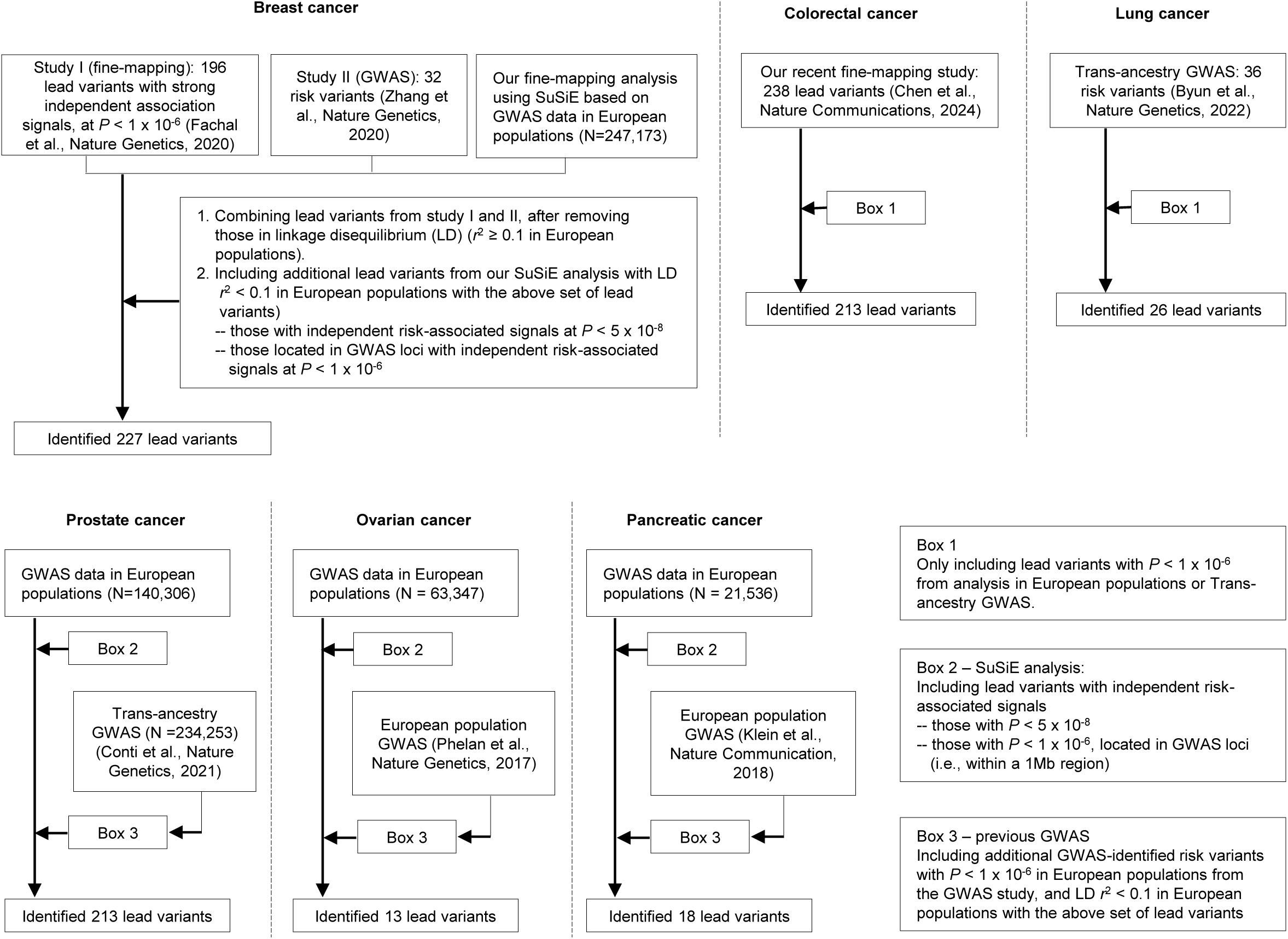
A flowchart for characterizing lead variants with independent risk signals in six types of cancer. The analysis for each of the six major cancers: breast, lung, colorectal, ovarian, pancreatic, and prostate is separated by dashed lines. The detailed protocols of new efforts from our additional fine-mapping analysis using SuSiE and a collection of previously identified risk variants from GWAS or fine-mapping studies are indicated in Box A, Box B and Box C, respectively.

**Figure S2:**
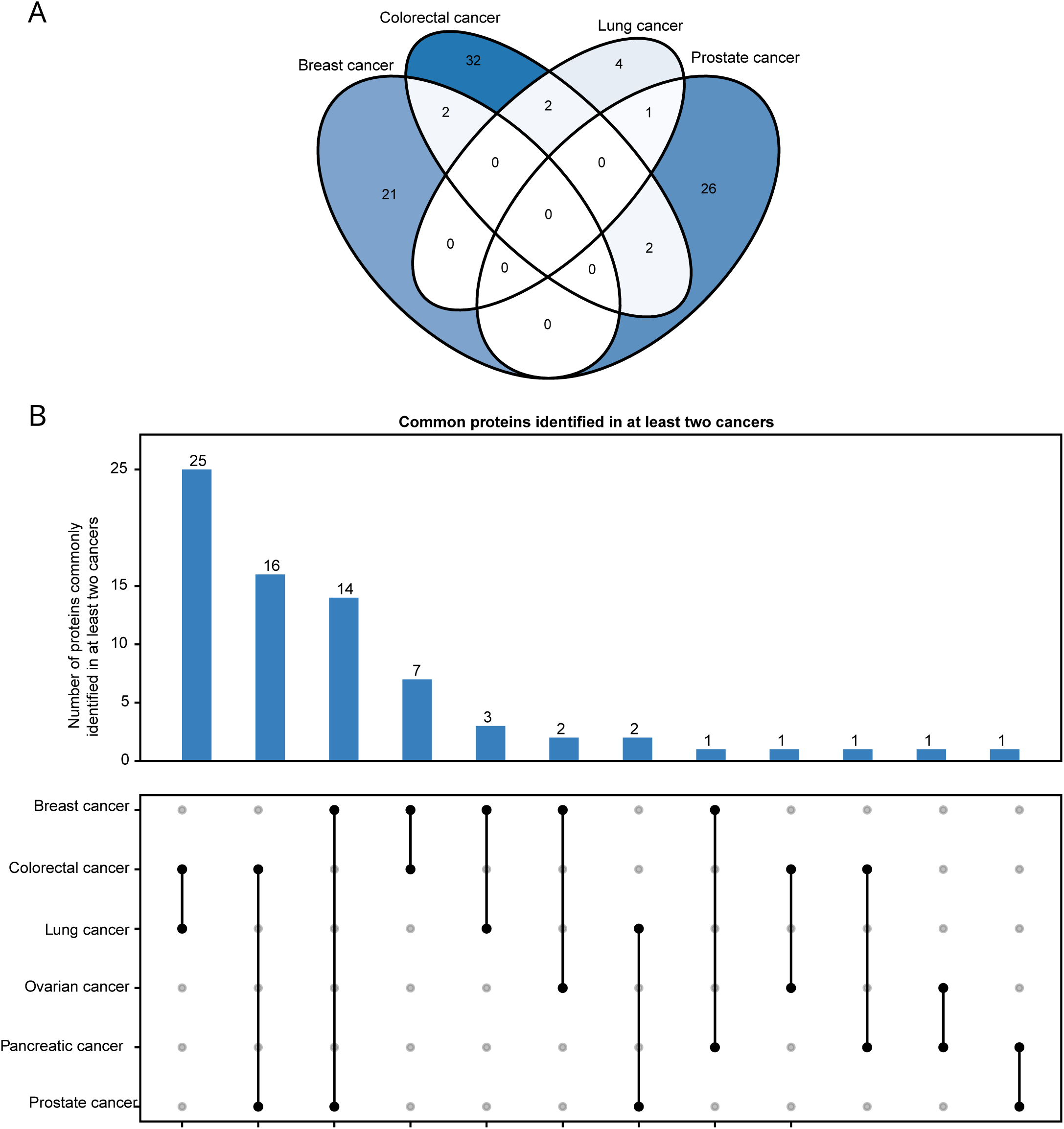
Common cancer risk proteins identified across six cancer types. **A**, A Venn plot showing common proteins in breast, colorectal, lung and prostate cancer. **B**, A heatmap showing common proteins observed from at least two of six types of cancer.

**Figure S3:**
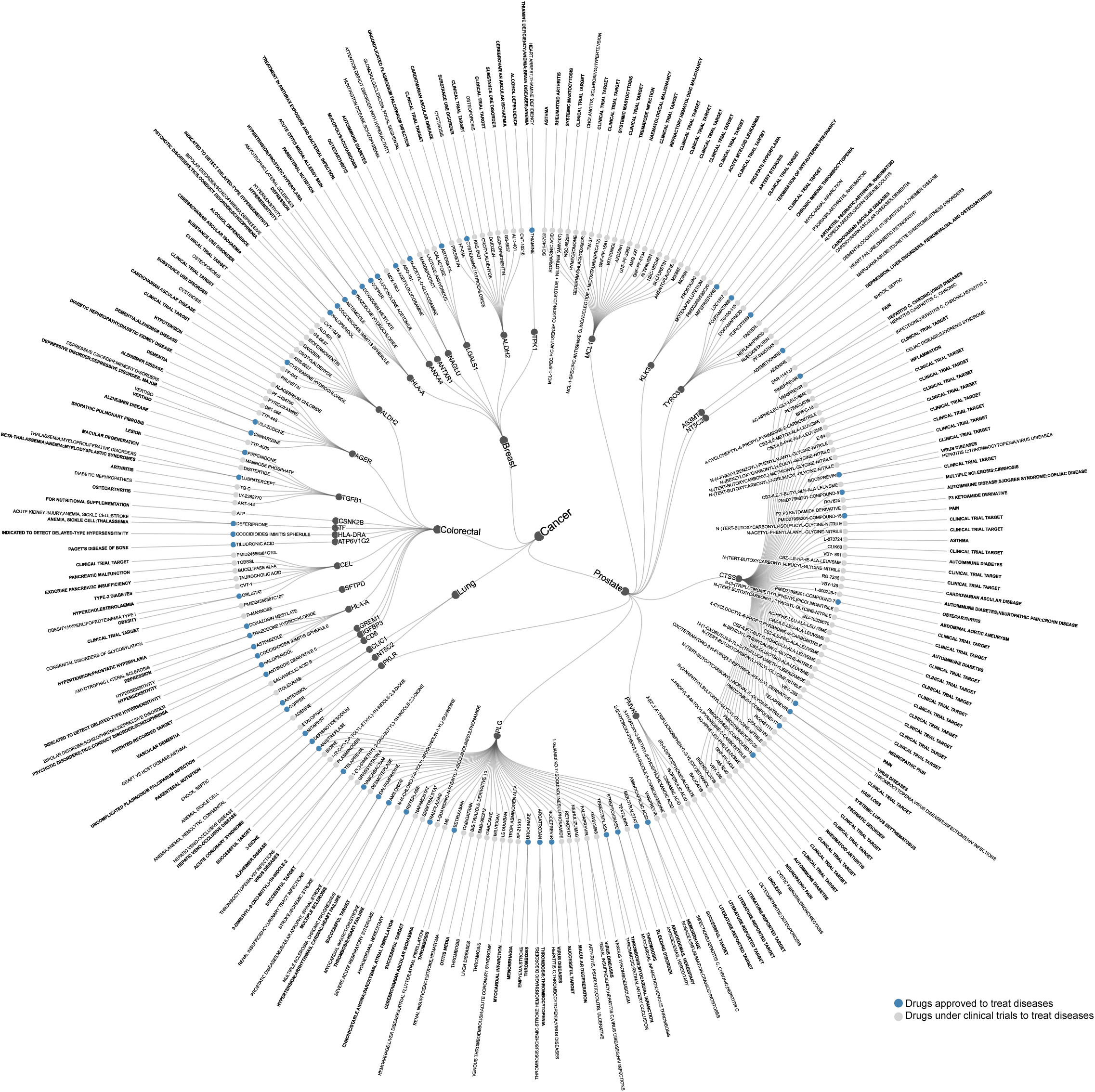
A circular plot showing 28 druggable proteins potentially targeted by 197 approved drugs or undergoing clinical trials for treated indications rather than cancers. Presented from inner to outer layers are cancer types, proteins, drugs and cancers. Drugs approved and undergoing clinical trials for cancer treatment are highlighted on blue and gray, respectively. Drug approved indications are formatted in bold, while indications under clinical trial are in regular font.

## Supplementary Tables

**Supplementary Table 1: GWAS summary statistics data of European descendants for breast, ovarian, colorectal, pancreatic, prostate and lung cancers used in this study.**

**Supplementary Table 2: List 710 lead variants with independent risk signals across six types of cancers**

**Supplementary Table 3: List of identified 365 putative cancer risk proteins across six types of cancers**

**Supplementary Table 4: List of novel and reported putative cancer risk proteins in breast, colorectal, ovarian, and prostate cancers**

**Supplementary Table 5: List of 101 cancer risk proteins identified by GWAS and pQTLs colocalization for six types of cancers**

**Supplementary Table 6: The identified cancer risk proteins supported by the evidence of eQTLs, sQTLs and apaQTLs**

**Supplementary Table 7: List of the identified cancer risk proteins potentially regulated by putative proximal regulatory variants.**

**Supplementary Table 8: List of the identified cancer risk proteins potentially regulated by putative distal regulatory variants.**

**Supplementary Table 9: Cancer risk proteins supported by the evidence of functional genomic analysis**

**Supplementary Table 10: List of 36 cancer druggable risk proteins Supplementary Table 11: List of cancer drugs and non-cancer drugs (for other indications) that potentially target cancer risk proteins**

**Supplementary Table 12: List of covariates utilized in LR-PS models**

**Supplementary Table 13: Individual Cox proportional hazards analysis results for treated and control drugs (10 years follow-up period)**

**Supplementary Table 14: Meta-analysis of Cox proportional hazards for treated vs. control drugs by level 2 ATC codes**

**Supplementary Table 15: Individual Cox proportional hazards analysis results for treated and control drugs (5 years follow-up period)**

**Supplementary Table 16: Meta-analysis of Cox proportional hazards for treated vs. control drugs by level 3 ATC codes**

## Notes

### Competing Interest Statement

The authors have declared no competing interest.

### Author Declarations

The study used ONLY openly available human data. The summary statistics of GWAS data for the six common cancers can be accessed through their publications, including Breast cancer(PMID: 32424353), Ovarian cancer(PMID: 28346442), Prostate cancer(PMID: 29892016), Colorectal Cancer(PMID: 30510241), Lung cancer(PMID: 28604730), Pancreatic cancer(PMID: 31917448). Metadata and pQTL summary statistics from UKB-PPP can be downloaded from Synapse: Project SynID: syn51364943; pQTL from ARIC46 and deCODE genetics47 can be accessed through previous publications (PMID: 34857953 and PMID: 35501419). Functional genomic data includes: TCGA and CPTAC differential expression results accessible through https://ualcan.path.uab.edu/index.html; 4DGenome: https://4dgenome.research.chop.edu/; Depmap：https://depmap.org/portal/; FANTOM5: http://fantom.gsc.riken.jp/5/. HaploReg: https://pubs.broadinstitute.org/mammals/haploreg/. GTEx: https://gtexportal.org/home/. GENCODE (v26.GRCh38) was downloaded from https://www.gencodegenes.org/human/release_26.html. National Cancer Institute can be accessed through https://www.cancer.gov/about-cancer/treatment/drugs; CGC can be accessed via COSMIC website: https://cancer.sanger.ac.uk/census. Drugs and compounds data can be downloaded from the following URLs: ChEMBL: https://www.ebi.ac.uk/chembl/; Therapeutic Target Database: https://db.idrblab.net/ttd/; Open Targets: https://www.opentargets.org/; DrugBank: https://go.drugbank.com/. The EHR data, containing de-identified clinical information, can be accessed through the VUMC SD database. Data is available through restricted access for approved studies and researchers who agree to specific conditions of use.

### Summary of Updates

Update manuscript, figures and tables

