## SupplementaryMaterials for "Large-scale integration of omics and electronic health records to identify potential risk protein biomarkers and therapeutic drugs for cancer prevention"

Dr. Xingyi Guo

Division of Epidemiology, Department of Medicine, Vanderbilt Epidemiology Center, and Vanderbilt-Ingram Cancer Center, Vanderbilt University School of Medicine

2525 West End Ave. Suite 330, Nashville, TN 37203

Dr. Zhijun Yin

Department of Biomedical Informatics, Department of Computer Science

Vanderbilt University Medical Center

2525 West End Ave. Suite 1475, Nashville, TN 37203

Dr. Quan Long

Department of Biochemistry and Molecular Biology,

University of Calgary,

3330 Hospital Drive NW

Room 1151 (1173), Health Sciences Centre

Calgary, AB T2N 4N1 Canada

**Supplementary Methods**

### Supplementary Method I. Data resources

The GWAS summary statistics data of European descendants for breast, prostate, ovarian, and lung cancers were downloaded and compiled from their corresponding consortia, including the Breast Cancer Association Consortium (BCAC)^1^ (N = 247,173; 133,384 cases and 113,789 controls), the Transdisciplinary Research of Cancer in Lung of the International Lung Cancer Consortium (TRICL-ILCCO) and the Lung Cancer Cohort Consortium (LC3)^2^ (N = 85,716; 29,266 cases and 56,450 controls), the Ovary Cancer Association Consortium (OCAC)^3^ (N = 63,347; 22,406 cases and 40,941 controls ), and the Pancreatic Cancer Case-Control Consortium (PanC4)^4^ (N = 21,536; 9,040 cases and 12,496 controls), and the Prostate Cancer Association Group Investigate Cancer Associated Alterations in the Genome (PRACTICAL)^5^ (N = 140,306; 79,194 cases and 61,112 controls). For colorectal cancer, we included GWAS data of 125,487 subjects from the European population.^6-8^ In addition, the GWAS data (N = 254,791) consisting of 100,204 colorectal cancer cases and 154,587 controls from European and Asian populations^9^ were also used in our analysis.

The large-scale *cis*-protein quantitative trait loci (*cis*-pQTLs) among European-ancestry populations were analyzed based on three proteomics datasets: UKB-PPP^10^ (N = 34,557; 2,922 plasma proteins), ARIC^11^ (N = 7,213; 4,657 plasma proteins) and deCODE^12^ genetics (N = 35,559; 4,907 plasma proteins). Detailed descriptions of sample collection and processes of the *cis*-pQTL analyses from the above proteomics datasets have been described in previous studies^10-12^. In brief, a 1 Mb window size for pQTLs (defined as variants within 1 Mb of the transcription start site) was used and results were extracted based on analyses conducted in the UKB-PPP (N = 34,557) and deCODE genetics (N = 35,559). Additionally, pQTL results using a 500 kb window size (N = 7,213) were extracted from analyses conducted in the ARIC study.

We utilized the synthetic derivative (SD), a database contains de-identified clinical information derived from Vanderbilt’s electronic medical records (EHRs) at Vanderbilt University Medical Center (VUMC)^13^. The SD has longitudinal clinical data for over 3.5 million individuals, including patient demographics, medical history, laboratory results, and medication history.

### Supplementary Method II. Inclusion criteria of patients for a focal drug and its control drugs

We enrolled patients for the treated group and the control group from the VUMC SD based on the following criteria: 1) patients aged ≥ 40 at the time of the latest EHRs or the initial diagnosis of cancer; 2) availability of at least one year of EHRs before the first prescription of the treated/control drug (index date); 3) for cancer patients, a minimum of two exposures to the treated or control drug, with at least a one-month gap, during the follow-up period (from the index date to the three months before cancer diagnosis, loss to follow-up, the end date of follow-up period (10 years), whichever came first); 4) for non-cancer individuals, a minimum of two exposures to control drugs, with at least a one-month gap, during the follow-up period (from the index date to the date of the latest EHRs, loss to follow-up, the end date of follow-up period (10 years), whichever came first). Finally, we precluded patients who were prescribed both treated and control drugs and discarded the trials with less than 500 eligible patients in each patient group^14^.

### Supplementary Method III. Emulation of treated-control drugs balanced trials

In the IPTW framework^15^, individuals are assigned weights based on the inverse of the probability of their received treatment, which represent their probability of being exposed to risk factors or a specific intervention, such as a treated drug, based on their baseline characteristics. In this study, we followed Zang’s work^14^ and trained a logistic regression propensity score (LR-PS) model with L2 regularization on patients' treatment assignments $Z$ and covariates. We followed VanderWeele^16^ and Brookhart et al^17^ by including covariates that are weakly associated with treatment assignment but strongly related to the outcome of interest (see Supplementary Table 12). We trained and selected the LR-PS model (Eq.1) using a 10-folder cross-validation (see Section VII). We used the selected model to calculate all patient’s stabilized weights (Eq. 2). These weights are used to calculate the standardized mean difference (SMD, Eq.3) of the covariate’s prevalence in treated and control groups. A covariate $d$ is defined as unbalanced if $SMD\left( d \right)>$ 0.1 in IPTW framework (Eqs. 3, 4). A trial is balanced if it contains $\leq$10% unbalanced covariates (Eq. 5).

The logistic regression is defined as follows:

| $\log\left( \frac{P\left( \boldsymbol{Z}=1 \right)}{1-P\left( \boldsymbol{Z}=1 \right)} \right)= \beta_{0}+\beta_{1}\boldsymbol{X}_{1}+\beta_{2}\boldsymbol{X}_{2}+\ldots+\beta_{n}\boldsymbol{X}_{n}$  $P\left( \boldsymbol{Z}=1 \right)= \frac{1}{1+ e^{-(\sum\beta_{j}\boldsymbol{X}_{j}+\beta_{0)}}}$ | 1 |
| --- | --- |

where $\boldsymbol{Z}$ refers to treatment assignment (1 for treated patient group and 0 for control patient group) and $\boldsymbol{X(}\boldsymbol{X}_{1}\boldsymbol{,}\boldsymbol{X}_{2}\boldsymbol{,}\boldsymbol{\ldots,X}_{n}\boldsymbol{)}$ for baseline covariates. The propensity score is defined as $P\left( \boldsymbol{Z}=1| \boldsymbol{X} \right)$ and the stabilized IPTW of each individual is calculated as follows:

| $\mathbf{w}=\frac{\boldsymbol{Z} \times P\left( \boldsymbol{Z}=1 \right)}{P\left( \boldsymbol{Z}=1\vert\boldsymbol{X} \right)} + \frac{\left( 1-\boldsymbol{Z} \right)\times(1-P\left( \boldsymbol{Z}=1 \right))}{1- P\left( \boldsymbol{Z}=1\vert\boldsymbol{X} \right)}$ | 2 |
| --- | --- |

We further truncate the $\mathbf{w}$ to the range of [$5\times{10}^{-6}, 50$], to mitigate the influence of the extreme values, which may introduce instable and bias to the calculatioin.

Standardized mean difference is calculated as following:

| $SMD\left( \boldsymbol{x}_{treat}-\boldsymbol{x}_{control} \right)= \frac{\left\vert\boldsymbol{\mu}_{treat}-\boldsymbol{\mu}_{control} \right\vert}{\sqrt{(\boldsymbol{S}_{treat}^{2}+ \boldsymbol{S}_{control}^{2})/2}}$ | 3 |
| --- | --- |

$\boldsymbol{x}_{treat}$, $\boldsymbol{x}_{control}\in R^{D}$ , representing vectors of $D$ number of covariates of treated group and control group respectively; $\boldsymbol{\mu}_{treat}$, $\boldsymbol{\mu}_{control}$ are their sample means, and $\boldsymbol{S}_{treat}^{2}, \boldsymbol{S}_{control}^{2}$ are their sample variances. In IPTW framework, the weighted sample mean $\boldsymbol{\mu}_{w}$ and sample variance $\boldsymbol{S}_{w}^{2}$are calculated as following:

| $\boldsymbol{\mu}_{w}= \frac{\sum\boldsymbol{w}_{i}\boldsymbol{x}_{i}}{\sum\boldsymbol{w}_{i}}$  $\boldsymbol{S}_{w}^{2}= \frac{\sum\boldsymbol{w}_{i}}{\left( \sum\boldsymbol{w}_{i} \right)^{2}-\sum\boldsymbol{w}_{i}^{2}}\sum\boldsymbol{w}_{i}{(\boldsymbol{x}_{i}- \boldsymbol{\mu}_{w})}^{2}$ | 4 |
| --- | --- |

Number of unbalanced covariates are calculated as following:

| $n=\sum_{d=1}^{D} \mathbb{1[}SMD(d)>0.1]$ | 5 |
| --- | --- |

$$where D is the total number of covariates in the model, d is one covariate$$

### Supplementary Method IV. Logistic regression propensity score (LR-PS) hyperparameter selection and model training

To select the optimal regulation penalty weight ($\lambda$), we applied 10-fold cross-validation on a list of lambda elements$\lambda\in$ [0.05, 0.1, 0.5, 1]. Specifically, the logistic model was trained on 9 training folders, and learnable parameters ($\beta$) were estimated through minimizing the binary cross-entropy loss with L2 penalty (Eq. 6). On the left-one out validation folder (k), we calculated ${SMD}_{k}$ (Eq. 3) values for $D$  covariates based on individuals’ weights (Eq. 2) as well as the number unbalanced covariates $n_{k}$ (Eq. 5). In addition, we evaluated trained model’s prediction performance using area under curve (${AUC}_{k}$) on the validation dataset. The same processes were repeated 10 times. We defined the optimal hyperparameter value is the value generates the smallest averaged $n_{k}$. For two hyperparameter values generate the same $n_{k}$, the one with larger averaged ${AUC}_{k}$ is the optimal. Finally, we trained LR-PS model’s learnable parameters ($\beta$) on all subjects with the optimal hyperparameter and leveraged this trained model to compute weights and the proportion of imbalanced covariates to pinpoint balanced trials.

Binary cross-entropy loss function with ridge (L2) penalization:

| ${argmin}_{\boldsymbol{\beta}}\left( Loss \right)=- \frac{1}{n}\left( \sum_{i=1}^{n} z_{i}\log\left( P\left( z_{i}=1 \right) \right)+\left( 1-z_{i} \right)\log\left( 1-P\left( z_{i}=1 \right) \right) \right)+ \lambda_{2}\sum_{j=1}^{b} \beta_{j}^{2}$ | 6 |
| --- | --- |

### Supplementary Method V. Calculation of overall hazard ratio for cancer risk drug on cancer development risk

We evaluate subjects’ hazard of developing/preventing cancer in balanced treated-control trials through survival analyses. We applied weighted Cox proportional hazards model^18^ with robust variance estimators using the lifelines 0.28.0 Python package to systematically evaluate the hazard ratio of developing cancer for patients taking treated drug vs patients taking control drugs (time-to-event). These weights were calculated through IPTW. The time windows utilized in this study start from the earliest date in EHRs for prescription of the treated/control drug to patients and end at the date of the diagnosis of cancer during follow-up period (event) or the end of EHRs records, lost follow-up, or end of follow-up period (censored). We included unbalanced covariates (if exist) into Cox models. For a treated drug, its overall hazard ratio and p-value were obtained by applying a random effect meta-analysis on the hazard ratios from its eligible trials (minimum of 10) using the meta 7.0 R package. We reported that a treated drug has a significantly increasing or decreasing risk of cancer development, contrasting to its control drugs, if the overall hazard ratio has a *P* < 0.05 after Bonferroni correction (nominal *P* = 4.5 x 10^-3^ corresponding to 11 tests).
